# Methylation-Based ctDNA for Post-Treatment Surveillance of Non-Viral Head and Neck Cancer

**DOI:** 10.64898/2026.09.17.26363361

**Authors:** Danielle Tomer, Patrik Brodin, Nikhil Malik, Sadiya Mayat, Christian Velten, Byung-Han Rhieu, Enrico Castellucci, Bradley A Schiff, Vikas Mehta, Richard V Smith, Shalom Kalnicki, Madhur Garg, Rafi Kabarriti

## Abstract

**Importance:** Recurrence rates in head and neck squamous cell carcinoma (HNSCC) remain high, yet current post-treatment surveillance relies primarily on clinical examination and imaging with known limitations. Circulating tumor DNA (ctDNA) has shown promise for recurrence detection in HPV-positive HNSCC, but less is known about its role in non-virally associated disease.

**Objective:** To evaluate the diagnostic performance of a methylation-based ctDNA assay for detecting recurrence during post-treatment surveillance of non-virally associated HNSCC and to assess its prognostic association with survival outcomes.

**Design, Setting, and Participants:** This retrospective single-institution cohort study included patients with non-metastatic non-virally associated HNSCC who underwent definitive therapy and had at least one *Guardant360* ctDNA test during post-treatment surveillance between January 2024 and October 2025. *Guardant360* identifies tumor-specific DNA methylation patterns via next-generation sequencing of plasma cell-free DNA. A positive result was defined by a methylation signal of 0.05% or greater.

**Main Outcomes and Measures:** Sensitivity, specificity, positive predictive value (PPV), negative predictive value (NPV), and area under the receiver operating characteristic curve (AUC). Progression-free survival (PFS) and overall survival (OS) were estimated using the Kaplan-Meier method.

**Results:** Fifty patients were included; most had larynx (38%) or oral cavity (36%) primaries, AJCC stage IVA–IVB disease (72%), T3–T4 tumors (69%), and N2–N3 nodal classification (48%). Treatment was chemoradiation (54%) or surgery (46%). At a median follow-up of 18.0 months, 13 patients recurred, and 12 had positive ctDNA results. Sensitivity, specificity, PPV, and NPV were 76.9%, 94.6%, 83.3%, and 92.1%, respectively (AUC, .858; 95% CI, .733–.982). Positive ctDNA was associated with worse PFS (1-year: 25.0% vs 96.4%; P < .001) and OS (1-year: 75.0% vs 100%; P < .001). Among 10 patients with recurrence and positive ctDNA, ctDNA preceded clinical detection in 6 (60%) by a median of 131 days. An exploratory analysis identified TP53, SETD2, and ROBO2 mutations as associated with recurrence.

**Conclusions and Relevance:** A methylation-based ctDNA assay demonstrated high sensitivity and specificity for detecting disease recurrence in non-virally associated HNSCC. Positive post-treatment ctDNA was associated with inferior survival and preceded clinical detection in most cases, supporting ctDNA integration into surveillance strategies, though prospective validation is warranted.

**Key Points:** *Question:* Can a methylation-based circulating tumor DNA assay detect recurrence during post-treatment surveillance of non-virally associated head and neck squamous cell carcinoma?

*Findings:* In this cohort study of 50 patients, the ctDNA assay had a sensitivity of 76.9% and specificity of 94.6% for detecting recurrence (AUC of 0.858). Positive ctDNA preceded clinical detection in 60% of recurrences by a median of 131 days and was associated with worse progression-free and overall survival.

*Meaning:* Methylation-based ctDNA may be a useful tool for surveillance of recurrence in non-virally associated HNSCC.

## Introduction

Despite advances in multi-modal treatment for head and neck squamous cell carcinoma, recurrences are prevalent, with more than 65% of patients with cancers of the head and neck developing recurrent or metastatic disease.^1^ The median time to recurrence diagnosis is still 5 to 40 months, with the majority of recurrences detected within two years.^2^ Current ASCO guidelines for surveillance include clinical visit follow-ups with the oncology team every few months, educating patients on the signs and symptoms of recurrence, and one-time post-treatment imaging within six months.^3^ Evidence on the benefits of imaging in surveillance is unclear, with some studies suggesting that imaging beyond six months may be useful in post-treatment surveillance,^4^ while others imply that imaging beyond two years may be costly, ineffective, and have no clear survival benefits.^5,6^

Recently, circulating tumor DNA (ctDNA) has been explored as a method of surveillance and recurrence detection of head and neck cancers, with particular sensitivity demonstrated in identifying residual disease in HPV-positive cancers.^7^ It has been previously shown that ctDNA is associated with high risk of tumor recurrence in breast, lung, and colorectal cancers.^8–10^ Liquid biopsies of blood, urine, and saliva have been used to detect circulating HPV DNA in patients with HPV-positive head and neck cancers.^11,12^ A review of studies on ctDNA in HPV-positive oropharyngeal cancers demonstrated that ctDNA levels correspond to disease burden and prognosis, that levels of clearance are associated with treatment response, and that post-treatment levels are associated with disease recurrence.^13^ In contrast, HPV-negative cancers are primarily driven by mutations that result from alcohol or tobacco use.^14^ However, ctDNA may be useful in surveillance of HPV-negative cancers too with data showing a recurrence detection rate of 80% and a median lead time of 4.6 months.^15^ However, the evidence in HPV negative cases is more limited when compared to HPV-driven disease, and more research is needed.^16^

The aim of this study was to investigate the utility of ctDNA in the post-treatment surveillance of HPV-negative non-metastatic head and neck squamous cell carcinoma (SCC) treated with definitive therapy, and to correlate ctDNA detection with disease status and survival.

## Methods

This retrospective clinical cohort study included sequential patients with non-virally associated head and neck SCC (HNSCC) treated at a single academic institution with at least one *Guardant360* ctDNA test during post-treatment clinical care and surveillance testing between January 2024 and October 2025. Inclusion criteria included patients with non-metastatic disease being treated with curative intent. Exclusion criteria included presence of metastatic disease, or presence of HPV- or EBV-associated disease. Guardant360 is an FDA-approved liquid biopsy assay that analyzes circulating tumor DNA (ctDNA) from plasma using next-generation sequencing to identify tumor-specific epigenomic alterations, including DNA methylation patterns.^17,18^ A positive ctDNA result in our study was defined by the observation of a tumor-specific methylation profile exceeding or equal to 0.05%. Clinical data and biomarker testing data were tabulated. ctDNA surveillance testing started at the end of definitive treatment. Clinical recurrence was defined by biopsy or clinical imaging. This study was approved by the Montefiore Medical Center Institutional Review Board, which waived the requirement for informed consent because the research involved retrospective review of existing clinical data with no more than minimal risk to participants.

### Statistical Analysis

Association between ctDNA positivity and patient demographics were assessed using chi-square tests for bivariate association for categorical variables and Wilcoxon rank-sum tests for continuous variables. Performance metrics for ctDNA testing were analyzed by computing the sensitivity, specificity, positive predictive value, negative predictive value and area under the receiver operating characteristics curve. Distribution of test results (false negatives, false positives, early true positives, and confirmatory true positives) and timing of events were summarized and visualized with a swimmer plot. Early true positives were defined as patients who had a positive ctDNA result that preceded recurrence, and confirmatory true positives were defined as positive ctDNA results that were concurrent with or shortly after a diagnosis of recurrence. Actuarial Kaplan-Meier survival estimates for overall and progression-free survival were generated to examine the prognostic value of ctDNA testing in this cohort. Sensitivity analyses were performed by stratifying Kaplan-Meier survival estimates by demographic or treatment variables shown to be associated (P < .10) with ctDNA positivity.

Mutations concurrently reported by the Guardant360 assay were tabulated by recurrence status. Genes with established associations with clonal hematopoiesis of indeterminate potential (CHIP), including DNMT3A, TET2, ASXL1, and PPM1D, were excluded from this analysis due to the absence of matched germline sequencing. Associations between individual gene mutation status and recurrence were assessed using Fisher’s exact test. Because this analysis was exploratory, P values were not adjusted for multiple comparisons.

## Results

Fifty patients met eligibility criteria with a median age of 64 years (range, 44 to 84 years). The most common HNSCC subsites were larynx (38%) and oral cavity (36%), with most patients having Stage IVA or IVB disease (72%). The median follow-up was 18 months (range, 4 to 80 months), during which 13 patients developed disease recurrence, of whom 6 died. A total of 62 ctDNA tests were performed in the cohort and the median time from treatment completion to ctDNA test was 8.7 months, with 12 patients having more than one ctDNA test performed.

A total of 12 patients (24%) had a positive ctDNA test with a median tumor fraction of 0.1% (IQR, 0.085% to 0.25%). Demographics are presented in Table 1. Other than age, baseline and clinical characteristics between ctDNA negative and ctDNA positive groups were comparable.

**Table 1.** Patient demographics.

|  | **ctDNA negative**<br>**(n=38)** | **ctDNA positive**<br>**(n=12)** | **P-value** |
| --- | --- | --- | --- |
| Sex, n (%) |  |  |  |
| Male | 27 (71%) | 5 (42%) | .064 |
| Female | 11 (29%) | 7 (58%) |  |
| Age, median (range) | 61 (44, 79) | 71 (60, 84) | .010 |
| Primary site |  |  |  |
| Larynx | 17 (45%) | 2 (17%) | .41 |
| Oral cavity | 13 (34%) | 5 (42%) |  |
| Oropharynx | 4 (11%) | 2 (17%) |  |
| Hypopharynx | 2 (5%) | 1 (8%) |  |
| Other | 2 (5%) | 2 (17%) |  |
| Staging, n (%) |  |  |  |
| Stage I-III | 10 (26%) | 4 (33%) | .64 |
| Stage IVa/IVb | 28 (74%) | 8 (67%) |  |
| Surgery, n (%) |  |  |  |
| Yes | 14 (37%) | 8 (67%) | .070 |
| No | 24 (63%) | 4 (33%) |  |

Of the 12 patients with a positive ctDNA test, 10 developed a recurrence while of the 38 patients with a negative ctDNA test, 3 developed a recurrence, corresponding to a sensitivity, specificity, positive predictive value (PPV), and negative predictive value (NPV) of 76.9% (95% CI, 65.2% to 88.6%), 94.6% (95% CI, 90.9% to 98.3%), 83.3% (95% CI, 72.5% to 94.1%), and 92.1% (95% CI, 87.7% to 96.5%), respectively. In terms of detecting recurrence, the area under the receiver operating characteristics curve (AUC ROC) for a positive ctDNA test was .858 (95% CI, .733 to .982). For the 10 patients with a documented recurrence and a positive ctDNA test, the ctDNA test was the first evidence of recurrence in 6 patients, with a median lead time of 131 days (range, 35 to 231 days). For the remaining 4 patients with documented recurrence, the positive ctDNA result was obtained shortly around the time of recurrence diagnosis. Figure 1 illustrates the distribution of early true positives, confirmatory true positives, as well as false positives and false negatives.

**Figure 1.**
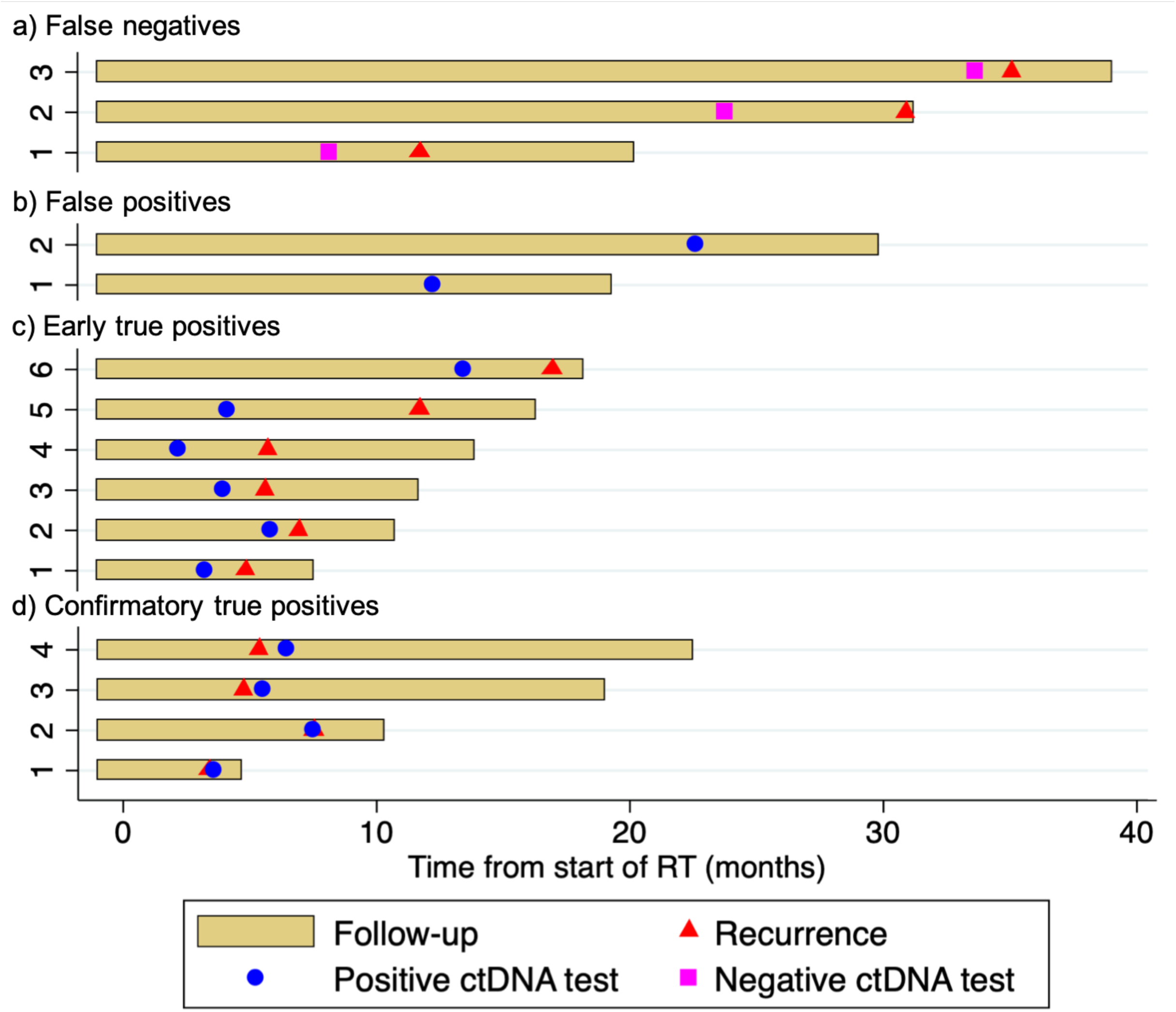
Swimmer plot showing distribution of false negatives, false positives, early true positives, and confirmatory true positives among 50 patients with non-virally associated HNSCC. Follow-up was defined from end of definitive treatment until last clinical assessment.

Progression-free survival (PFS) was significantly worse for patients who were ctDNA positive vs negative with 1-year PFS 25.0% vs 96.4% (log-rank P < .001). Similarly, 1-year overall survival from end of definitive treatment was 75.0% vs 100% for patients who were ctDNA positive vs negative (log-rank P < .001). Figure 2 shows the actuarial overall and progression-free survival for patients with and without positive ctDNA test.

**Figure 2.**
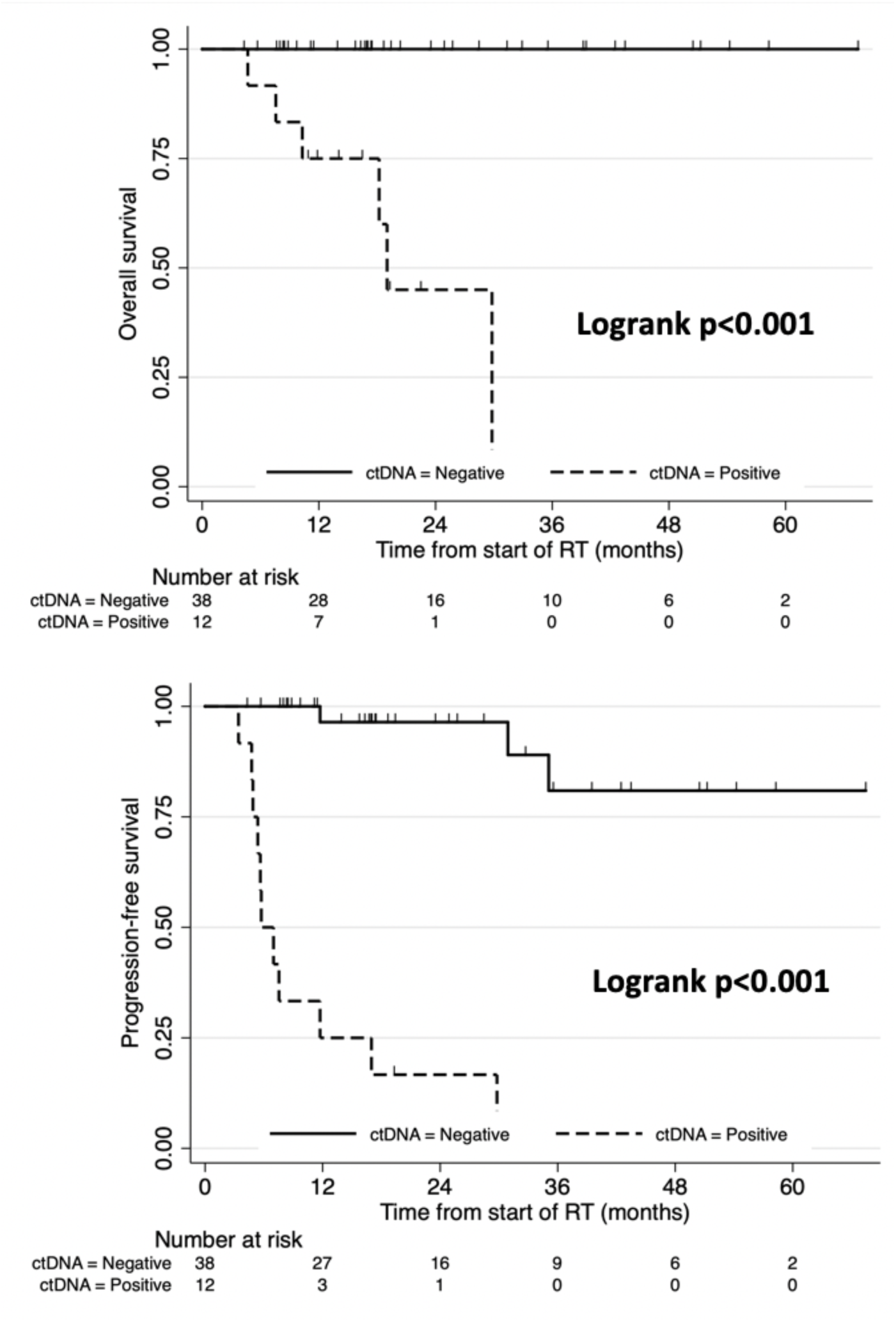
Kaplan-Meier survival curves showing overall survival and progression-free survival for patients with and without a positive ctDNA test (N = 50). Log-rank P < .001 for both comparisons.

The limited number of patients and events does not provide sufficient statistical power for multivariable analyses, but stratification was performed as a sensitivity analysis for variables associated with ctDNA positivity (sex, age and surgery). The comparisons shown in Table 2 illustrate that ctDNA positivity was strongly associated with survival outcome regardless of age, sex or whether the patients had surgery.

**Table 2.** Kaplan-Meier survival estimate Logrank tests for ctDNA positivity, stratified by associated patient demographic and treatment variables.

|  | **Overall survival** | **Progression-free survival** |
| --- | --- | --- |
| Sex |  |  |
| Male | P < .001 | P < .001 |
| Female | P = .003 | P < .001 |
| Age |  |  |
| ≥ median (61 y) | P < .001 | P < .001 |
| < median (61 y) | N/A* | P = .001 |
| Surgery, n (%) |  |  |
| Yes | P = .005 | P < .001 |
| No | P < .001 | P < .001 |

\*No deaths in the age < median group

In addition to the methylation-based ctDNA signal, the Guardant360 assay also reports mutations identified through targeted sequencing of cancer-associated genes. After exclusion of putative CHIP variants (DNMT3A, TET2, ASXL1, PPM1D), an exploratory analysis of mutations by recurrence status was performed (eTable 1 in Supplement). TP53 was the most frequently mutated gene overall (21/50, 42.0%) and was detected at a significantly higher rate in patients who recurred compared with those who did not (69.2% vs 32.4%; P = .027). Two additional genes, SETD2 and ROBO2, were significantly associated with recurrence (23.1% vs 2.7%; P = .049 for both). Several other HNSCC driver genes, including PIK3CA, EGFR, NOTCH1, and CDKN2A, showed higher mutation frequencies in the recurrence group but did not reach statistical significance. These gene-level comparisons are exploratory and were not adjusted for multiple comparisons given the small sample size.

## Discussion

Circulating tumor DNA is a promising biomarker for post-treatment surveillance in head and neck cancer. However, its clinical utility in predicting recurrence of head and neck cancer, especially EBV and HPV-negative subtypes, remains under investigation. This retrospective study showed that ctDNA can predict cancer recurrence in non-virally mediated head and neck cancers with high sensitivity and specificity, making it a clinically useful test. Moreover, positive ctDNA results were the first evidence of cancer recurrence in most patients with documented recurrence, and patients with ctDNA detection after cancer treatment had worse survival outcomes compared to patients who had negative ctDNA results. Therefore, our data strongly suggest that the use of ctDNA may be helpful in detecting minimal residual disease and diagnosing microscopic recurrences before progression.

ctDNA demonstrated high specificity (94.6%) and moderately high sensitivity (76.9%) in this cohort, suggesting that patients who test positive during post-treatment surveillance are very likely to have a recurrence. These findings are consistent with studies showing that ctDNA is a promising biomarker in patients with HPV-positive head and neck cancer. In the primary diagnosis setting, one study showed serum HPV ctDNA to have a sensitivity of 98.4% in initial diagnosis of HPV-positive head and neck cancers and a shortened time to diagnosis by 26 days.^19^ In addition, our results are similar to the findings in a study of colorectal cancer patients, which found that ctDNA had a sensitivity of 83.3% and specificity of 89.5% at predicting tumor recurrence.^17^ In HPV positive oropharyngeal squamous cell carcinoma, Chera et al demonstrated that two consecutive positive tests had a PPV of 100%, and median lead time between HPV DNA positive and biopsy-proven recurrence was 6.6 months.^20^ While the use of ctDNA for surveillance is well established in HPV positive head and neck cancers, studies evaluating ctDNA in HPV negative cancer–particularly in the post-treatment surveillance setting–remain limited; our findings contribute to emerging evidence supporting its reliability for recurrence detection.

Furthermore, ctDNA was the first evidence of recurrence in most patients with a documented recurrence in our study, and ctDNA may offer greater sensitivity and specificity for detecting recurrences than conventional imaging modalities. Indeed, one study showed that CT scans had a sensitivity and specificity of 63% and 80% respectively for detecting head and neck cancers in patients with suspected recurrence,^21^ which has lesser accuracy than ctDNA when compared with our results. Similar findings have been demonstrated in patients with non-small cell lung cancer, with one study showing that ctDNA positivity precedes radiologic recurrence by a median of 88 days.^22^ This suggests that ctDNA may identify recurrences at an earlier timepoint and when the disease burden is less compared to other modalities. Another study in head and neck cancers found that post-treatment ctDNA had the same sensitivity as PET imaging for detecting recurrence, but ctDNA had a higher specificity than PET imaging, suggesting ctDNA may be better at detecting true positives compared to PET scans.^23^ The higher specificity of ctDNA may also reduce unnecessary patient anxiety from false-positive imaging results. Taken together, these findings suggest that ctDNA, in addition to being simpler and less costly, may be a more clinically useful tool for post-treatment surveillance.

Patients with positive ctDNA also had significantly worse progression-free and overall survival. This is consistent with findings by Hanna et al, which demonstrated that HPV-negative head and neck cancer patients with post-treatment ctDNA positivity had significantly worse progression-free survival.^24^ Interestingly, a meta-analysis of ctDNA in HPV-negative cancers identified an association of specific ctDNA mutations in the TP53 and DNA repair genes and DNA methylation changes in SEPT9/SHOX2 genes with worsened survival outcomes in HPV-negative cancer.^25^ Identifying specific molecular biomarkers through ctDNA may further refine its efficacy as a surveillance tool for recurrences. Notably, Chen et al demonstrated that surveillance imaging with PET/CT, MRI, or CT alone did not improve local-regional control, survival, or distant metastases development compared with patients who received expectant management.^26^ Further studies are needed to determine if surveillance with ctDNA improves outcomes compared to other methods of surveillance.

An exploratory analysis of mutations reported by the Guardant360 assay showed that TP53 mutations were more frequent in patients who recurred (69.2% vs 32.4%; P = .027). TP53 ctDNA mutations have also been associated with reduced overall survival, disease-free survival, and progression-free survival in HPV-negative head and neck cancer.^25^ As the most frequently mutated gene in HPV-negative HNSCC, with loss-of-function mutations occurring in the majority of cases,^27^ TP53 mutations have been independently associated with increased risk of locoregional recurrence.^28^ In contrast, SETD2 and ROBO2 have not been previously identified as recurrence-associated genes in HNSCC ctDNA surveillance studies. SETD2 is a histone methyltransferase with tumor-suppressive function across multiple solid tumor types,^29^ and ROBO2 inactivation has been described in early dysplastic head and neck lesions but not in the post-treatment ctDNA setting.^30^ If validated in larger cohorts, these associations could help refine risk stratification for recurrence, and the ability to detect both methylation signals and mutations from a single blood draw makes this a practical approach. However, without matched germline sequencing, CHIP-associated variants cannot be definitively excluded. Prospective studies incorporating paired tumor and germline sequencing are needed to validate these findings and distinguish tumor-derived variants from potential non-tumor sources.

Our study has several limitations. Our small sample size limits the statistical power of our analysis. Being conducted at a single academic center, it is possible that our results may lack generalizability to other populations, but it is an important study as our population is unique and very diverse. More multi-center studies are needed to determine the utility of ctDNA across broader populations. Finally, it is possible that recurrences are underestimated due to the limited time for follow-up, and that our results are mostly applicable for early recurrences.

## Conclusions

Recurrences in head and neck cancers are common and the efficacy of post-treatment surveillance with imaging alone has yielded mixed results. ctDNA has the potential to be a sensitive and specific method for detecting minimal residual disease and recurrences of head and neck cancer treated with definitive therapy.

## Data Availability

The de-identified data that support the findings of this study are available from the corresponding author upon reasonable request.

## Author Contributions

Dr. Kabarriti had full access to all the data in the study and takes responsibility for the integrity of the data and the accuracy of the data analysis.

Concept and design: All authors

Acquisition, analysis, or interpretation of data: All authors

Drafting of the manuscript: All authors

Critical review of the manuscript for important intellectual content: All authors

Administrative, technical, or material support: Rafi Kabarriti

Supervision: Rafi Kabarriti

## Conflict of Interest Disclosures

None

## Funding/Support

None

## Role of the Funder/Sponsor

Not applicable

---

**eTable 1.**
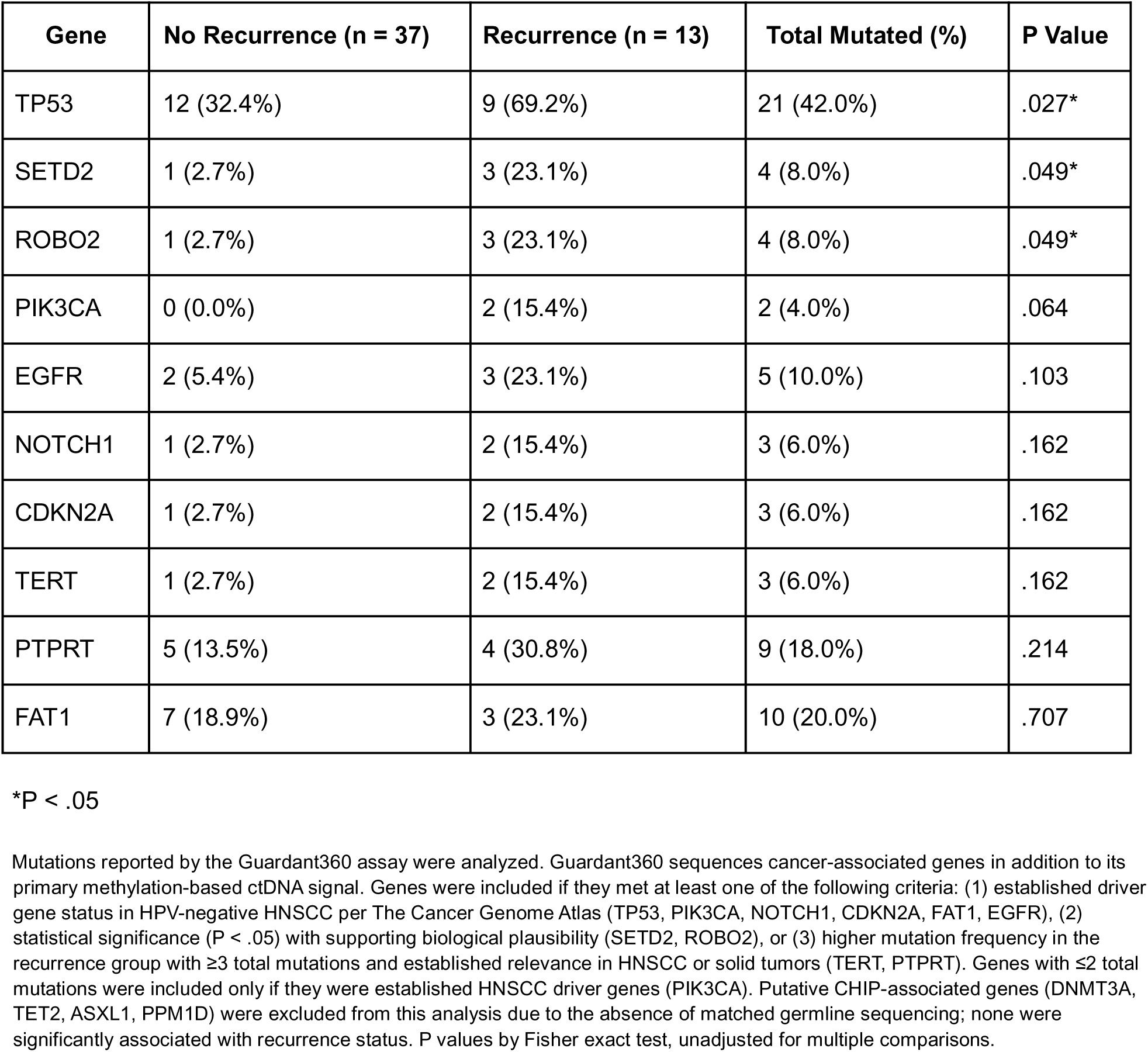
Mutations Detected by Guardant360 According to Recurrence Status in Patients With HNSCC (N = 50)

## Notes

### Competing Interest Statement

The authors have declared no competing interest.

### Author Declarations

The IRB of Albert Einstein College of Medicine gave ethical approval for this work.

